# Appraisal of the causal effect of *Chlamydia trachomatis* infection on epithelial ovarian cancer risk: a two-sample Mendelian randomisation study

**DOI:** 10.1101/2024.10.13.24315417

**Authors:** Sarah L. Perrott, Siddhartha P. Kar

## Abstract

**Background:** History of *Chlamydia trachomatis* infection has previously been associated with epithelial ovarian cancer (EOC) in observational studies. We conducted a two-sample univariable Mendelian randomisation (MR) study to examine whether genetically predicted seropositivity to the *C. trachomatis* major outer membrane protein (momp) D is causally associated with EOC.

**Methods:** MR analyses employed genetic associations derived from UK Biobank as proxies for momp D seropositivity in 25 509 EOC cases and 40 941 controls that participated in the Ovarian Cancer Association Consortium. Findings were replicated using a GWAS meta-analyses of global biobanks including the UK Biobank, FinnGen and BioBank Japan.

**Results:** Genetically predicted momp D seropositivity was associated with overall and high-grade serous EOC risk in inverse-variance weighted (IVW) and MR-Egger univariable MR analysis (odds ratio (OR) 1.06; 95% confidence interval (CI) 1.02—1.10, and OR 1.08; 95%CI 1.01—1.16, respectively). Replication yielded similar results for overall EOC (OR 1.11; 95%CI 1.01—1.22).

**Conclusion:** This MR study supports a causative link between *C. trachomatis* infection and overall and high-grade serous EOC.

## Introduction

*Chlamydia trachomatis* is the most prevalent sexually transmitted infection (STI), affecting 4% of women worldwide.^1^ *C. trachomatis* infections are asymptomatic in up to 70%, and consequently often go undiagnosed and untreated, resulting in onward transmission and sequelae including pelvic inflammatory disease (PID) and infertility in women.^2^ Both *C. trachomatis* and PID have been associated with an increased risk of ovarian cancer, the most lethal gynaecological cancer.^3–5^

Ovarian cancer is a global problem; although central Europe has the highest incidence, rates in low-and middle-income countries have increased by approximately 70% since 2007.^6^ Traditional risk factors include increased age, inherited germline mutations, family history, and infertility treatments. Parity, lactation and oral contraceptives are recognised as protective factors.^7^ Ovarian tumours originate from either epithelial cells (90%), stromal cells (5-6%), or germ cells (2-3%). Epithelial ovarian cancer (EOC) is a heterogeneous group and can be further subdivided by histotype into high grade serous; low grade serous; mucinous; clear cell; and endometrioid EOC. High-grade serous EOC is not only the most common, accounting for 75% of all ovarian cancers, but also the most aggressive and deadly.^8^ The conventional EOC risk factors, once accepted for all subtypes, are strongly associated with non-serous rather than serous EOCs.^7^ Recent molecular and epidemiological evidence suggest that high-grade serous EOCs do not even originate from the ovary, but precancerous lesions of the fimbriated end of the fallopian tubes called serous tubal intraepithelial carcinomas (STICs).^9^ STICs have previously been associated with pelvic inflammatory conditions including endometriosis and PID.^10^

Fallopian tube inflammation and damage are often secondary to PID, prompting consideration of the potential involvement of infectious agents in both STICs and EOC. The relationship between pathogenic organisms and cancer is well-established; helicobacter pylori has been long associated with gastric cancer and human papilloma virus (HPV) with cervical cancer.^11,12^ The associations between PID and EOC, and several infectious agents including *C. trachomatis*, *Neisseria gonorrhoeae*, *Mycoplasma genitalium*, HPV, and herpes simplex virus 2 (HSV-2) have been previously examined in observational studies, namely case-control or prospective cohort.^3–5,13–18^ When considered overall, evidence supporting the role of *C. trachomatis* in tubal inflammation, damage, and potentially ovarian carcinogenesis, is more compelling compared to that of other STIs.^7,19^ Despite this, a few studies have failed to demonstrate any association, or have suggested HSV-2 or *M. genitalium* may have a role instead.^5,14,16–18^ The existing evidence base is deficient due to challenges in study design, influenced by residual confounding factors and relatively small study populations. *C. trachomatis* is a small obligate intracellular bacterium that has developed mechanisms to promote bacterial survival, which potentially lead to host cell genomic instability, cell cycle disruption, and inhibition of apoptosis – all of which are hypothetical hallmarks of a carcinogenic pathogen.^10^ However, it has not been possible to determine whether observed associations are causal, or whether other external factors influence future EOC risk.

Mendelian randomisation (MR) is an epidemiological strategy aimed at removing potential biases which exist within conventional observational studies.^20^ This method uses single nucleotide polymorphisms (SNPs) as instrumental variables, enabling a potential causal relationship between an exposure and outcome to be determined. To our knowledge, MR has never been used to explore this association. We therefore used a two-sample univariable MR approach to investigate the causal relationship between seropositivity to the *C. trachomatis* major outer membrane protein (momp) D antibody and EOC. By elucidating the causal relationship between *C. trachomatis* seropositivity and EOC, this study aims to contribute to our understanding of EOC aetiology – particularly with regard to serous EOC, whereby any modifiable risk factors yet remain undetermined.^7^

## Methods

### Study design

The current study has been reported according to the STROBE-MR guidelines (Reporting Standards Document). This study employed a two-sample univariable MR design to investigate the causal relationship between *C. trachomatis* seropositivity and EOC. To establish causality, MR relies on three key assumptions as illustrated in Figure 1.

**Figure 1:**
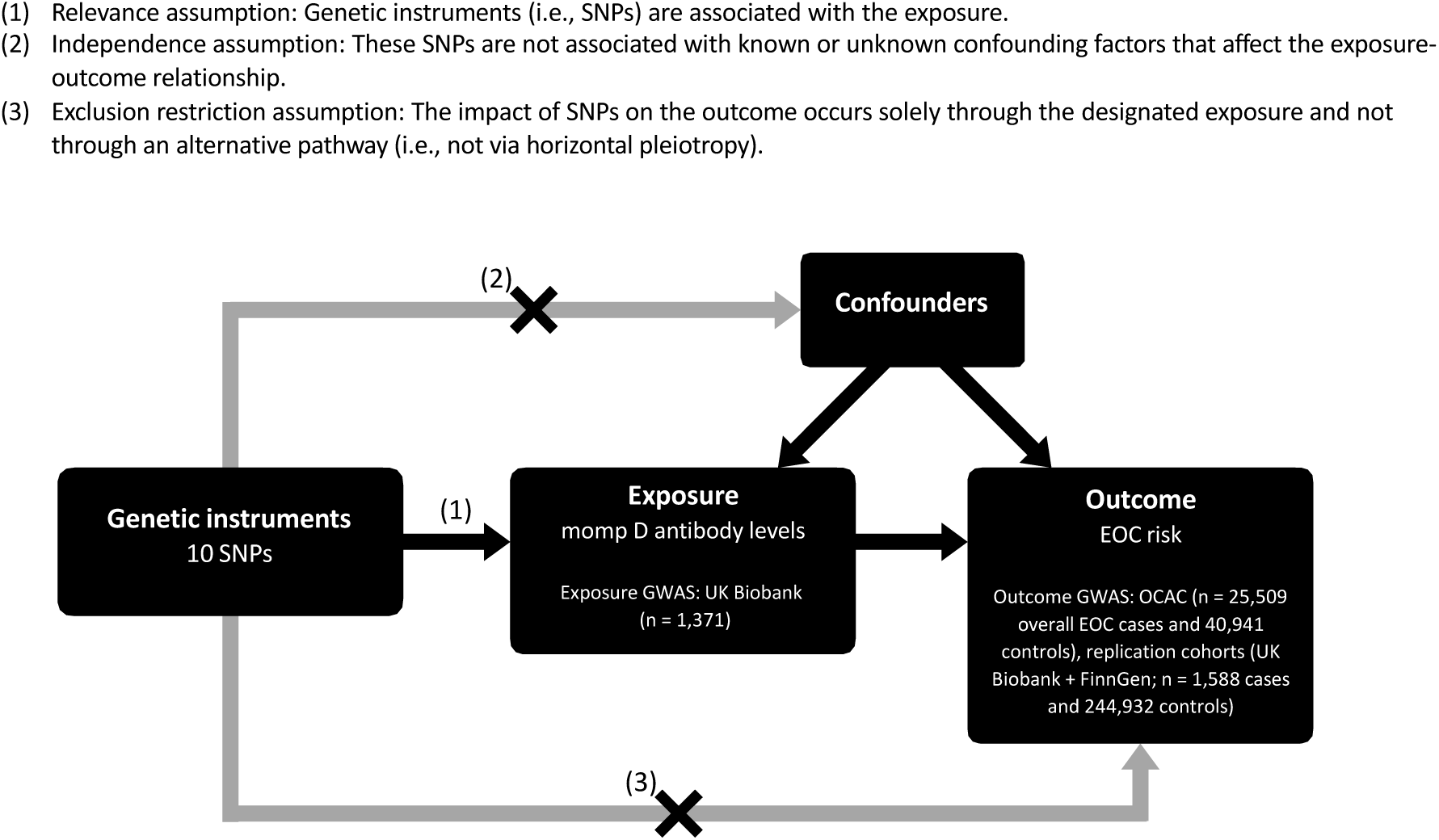
Study design and assumptions of two-sample Mendelian randomisation analysis.

The MR approach utilises SNPs as instrumental variables for the exposure, allowing the lifelong and causal effects of the exposure on the outcome to be examined. Since SNPs are randomly assigned at conception, an MR study theoretically mirrors the design of a randomised controlled trial, minimising effects of confounding through randomisation. Compared to conventional epidemiological studies, this makes MR studies less susceptible to reverse causation and unmeasured confounding, therefore providing stronger evidence of causal inference. This study uses the two-sample MR approach; this means summary statistics were obtained for the exposure and outcome from two independent genome-wide association study (GWAS) samples. In all analyses, we used univariable MR approach to evaluate if genetic liability to *C. trachomatis* phenotypes were causally associated with EOC. We utilised previously validated sets of single nucleotide polymorphisms (SNPs) associated at genome-wide significance (*p* < 5x10^-8^) with *C. trachomatis* momp D seropositivity and EOC (overall, and by histotype). Additionally, we conducted several supplementary analyses to assess the robustness of our MR results, including sensitivity analyses and assessments for potential horizontal pleiotropy (i.e. ensuring SNPs influence the outcome only through the allocated exposure, and not through any other pathway). Publicly available summary statistics were retrieved from GWAS conducted predominantly in Europeans. All included GWASs and biobanks were approved by corresponding ethics committees and participants provided written informed consent.

#### C. trachomatis serology (exposure) data

The exposure evaluated was *C. trachomatis* seropositivity level based on the serum momp D antibody. Momp D was chosen over other serological markers as there were multiple genome-wide significant (*p* < 5 x 10^-8^) SNPs associated with it, allowing for the use of the powerful inverse-variance weighted MR method and for robust sensitivity analyses to evaluate the exclusion restriction assumption of MR (Figure 1). We used genome-wide significant SNPs associated with serum momp D antibody level in the UK Biobank from a GWAS by Butler-Laporte *et al*.^21^ The UK Biobank is a prospective cohort study that recruited approximately 500,000 individuals of predominantly European ancestry, aged 40 to 69 years, at 22 assessment centres across England, Scotland, and Wales during 2006–2010. Of these, a subset of 9,724 individuals provided serum for serological evaluation of 20 common infectious microorganisms based on fluorescent bead multiplex technology. Butler-Laporte *et al* undertook GWAS for infections with a seroprevalence of over 15% in this subset of the UK Biobank, performing genetic association analyses on log-transformed mean fluorescence intensity (MFI) levels (a standardised measure of antibody level) in European ancestry individuals. A total of 1,371 individuals were seropositive for momp D (defined as MFI > 100) and included in the momp D GWAS. Ten independent *p* < 5 x 10^-8^ SNPs from the momp D GWAS (linkage disequilibrium *r2* = 0.001 and clumping distance = 10,000 kilobase pairs used to defined independence) were employed collectively as a genetic instrument for momp D seropositivity level.

#### Epithelial ovarian cancer risk (outcome) data

To evaluate the association between genetically predicted serum momp D level and risk of overall and histological subtype-specific EOC, we obtained summary genetic association data from a GWAS meta-analysis by Phelan *et al* and the Ovarian Cancer Association Consortium (OCAC).^22^ This GWAS meta-analysis included 25,509 EOC cases and 40,941 controls. Associations were reported for EOC overall (n = 25,509 cases), and for seven histotypes: high-grade serous (n = 13,037 cases), low-grade serous (n = 1,012 cases), invasive mucinous (n = 1,417 cases), clear cell (n = 1,366 cases), endometrioid (n = 2,810 cases), serous low malignant potential (n = 1,954 cases), and mucinous low malignant potential (n = 1,149 cases), with each histotype compared against 40,941 controls. The GWAS meta-analysis was based on individuals of genetically inferred European ancestry.^23,24^ Given the large sample size of this OCAC GWAS meta-analysis and inclusion of major EOC histotypes, these outcome data were used for the MR primary analyses.

Findings were replicated using a GWAS meta-analysis of two independent biobanks, the UK Biobank and FinnGen, that together included 1,588 EOC cases and 244,932 controls of genetically inferred European ancestry.^25^ This study by Sakaue *et al* combined genotype and phenotype data from these global biobanks to identify genetic associations with EOC risk (not subdivided by histotype). Disease endpoint data were derived from electronic medical records and health registries in the two biobanks. This GWAS meta-analysis provided a distinct, independent population that was ideal for replication purposes.

### Statistical analyses

Analyses were performed using the *TwoSampleMR* package on R version 4.3.2.20 This package facilitates semi-automated data formatting, harmonisation, and application of MR methods.

Genetic associations for EOC from the OCAC GWAS were imputed using the *1000 Genomes Project* reference panel. ^22^ The global biobanks GWAS data were imputed using a combination of reference panels, including the *Haplotype Reference Consortium*, *UK10K*, *1000 Genomes Project*, and a Finnish-specific whole genome sequenced reference panel.^26^ Linkage disequilibrium clumping was performed to ensure instruments for the exposure are independent, removing one correlated SNP (rs140290448). Summary statistics for the 10 remaining momp D-associated lead SNPs were available in both EOC data sets (OCAC and global biobanks). These exposure and outcome summary statistics were then subject to allele harmonisation by aligning the beta-coefficients to ensure that the effect allele in the exposure data set is the same effect allele in the outcome data set. For rs140290448, no proxy SNPs in linkage disequilibrium were identified.

For the primary analysis, effect estimates for EOC association by histotype with momp D seropositivity were generated using the inverse-variance weighted median (IVW) method with random-effects using the OCAC GWAS data. The IVW method is widely used in MR; it combines SNP-exposure and SNP-outcome associations using inverse-variance weights, maximising statistical power of the analysis – particularly when multiple instrumental variables are available. It is consistent and easily interpreted, however can be sensitive to outliers and horizontal pleiotropy. Therefore, we conducted four sensitivity analysis (MR-Egger, weighted-mean, simple mode, and weighted mode) to examine the consistency of results and correct for any directional pleiotropy. These alternative MR methods are more robust to pleiotropic variants and were performed for overall EOC and high-grade serous ovarian cancer. We also performed the MR-Egger intercept; this can indicate any potential pleiotropic effects of the genetic instruments.

For the secondary (replication) analysis, overall EOC associations with momp D seropositivity was investigated using the global biobanks dataset. Finally, meta-analysis of effect estimates (both by IVW method) from both OCAC and global biobanks were performed, providing an overall effect estimate. The strength of the 10-SNPs was also evaluated using the F-statistic, with F<10 considered an indicator of a weak instrument.^27^ All statistical tests were 2-sided and *p*<0.05 was considered statistically significant unless otherwise specified. Odds ratios (OR) and 95% confidence intervals (95% CI) were presented as forest plots using the *ggplot2* R package, version 4.3.2. Finally, this study has been reported as per the Strengthening the Reporting of Observational Studies in Epidemiology (STROBE)-MR guideline.^28^

## Results

Associations between individual SNPs and the risk of EOC overall and by histotype in the OCAC and in the combined UK Biobank and FinnGen GWAS meta-analyses are provided in Supplementary Tables 1 and 2, respectively. Each SNP was associated at genome-wide significance (p < 5 x 10^-8^) with momp D antibody seropositivity level and collectively, they yielded an F-statistic of 112, indicating adequate instrument strength given that F was > 10.

### Primary Mendelian randomisation and sensitivity analyses

Genetically predicted elevated momp D antibody seropositivity level was associated with increased risk of EOC overall and EOC of the high-grade serous histotype in the primary inverse variance weighted MR analysis (overall EOC OR 1.06 (95% CI 1.02–1.10; *p* = 0.004; Figure 2; all odds ratios and confidence intervals are in units of per standard deviation increase in log-transformed antibody mean fluorescence intensity), and high-grade serous EOC OR 1.08 (95% CI 1.01–1.16; *p* = 0.04; Figure 2)). The point estimate of the effect size for the association with low-grade serous EOC risk was consistent with the high-grade serous EOC association (1.10 (95% CI 0.91–1.31; *p* = 0.32; Figure 2)). There was no evidence of an association with risk of invasive mucinous, clear cell, endometrioid, serous low malignant potential, and mucinous low malignant potential EOC (Figure 2). The inverse variance weighted estimates for risk of overall EOC and high-grade serous EOC were consistent in direction with the results of the MR-Egger and the weighted median methods in sensitivity analyses (Figure 3). Sensitivity analyses results for all histotypes are provided in Supplementary Table 3. The MR-Egger intercept test (*p* = 0.25 for both overall and high-grade serous EOC) suggested absence of directional horizontal pleiotropy (i.e., it provided statistical evidence for the absence of an effect of the 10 SNPs collectively on overall or high-grade serous EOC risks via a pathway other than momp D serology; Supplementary Table 4).

**Figure 2:**
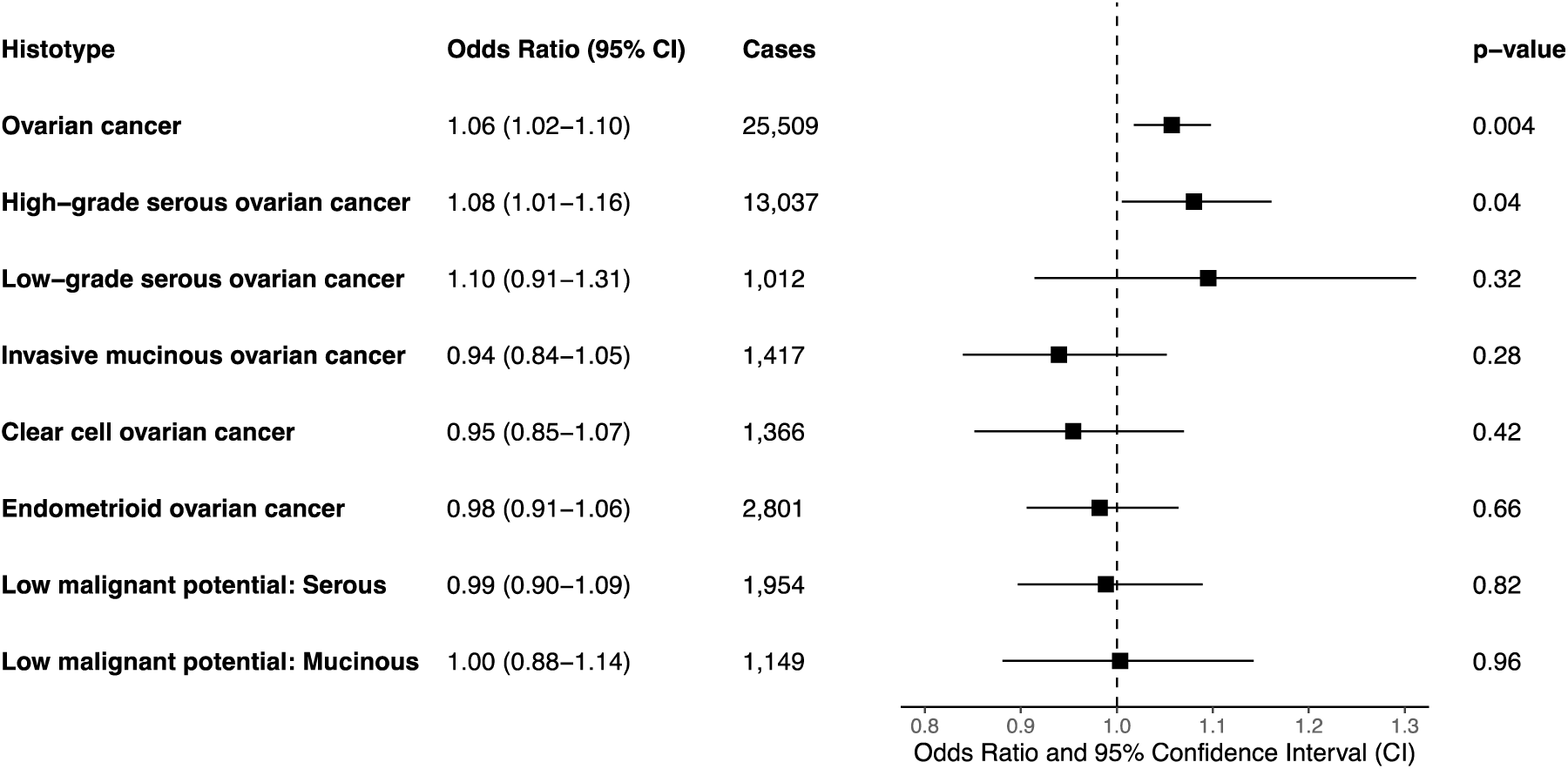
Inverse variance weighted Mendelian randomisation analysis results for the association between genetically predicted momp D antibody seropositivity level and overall and histotype-specific epithelial ovarian cancer risk. All odds ratios and confidence intervals are in units of per standard deviation increase in log-transformed antibody mean fluorescence intensity.

**Figure 3:**
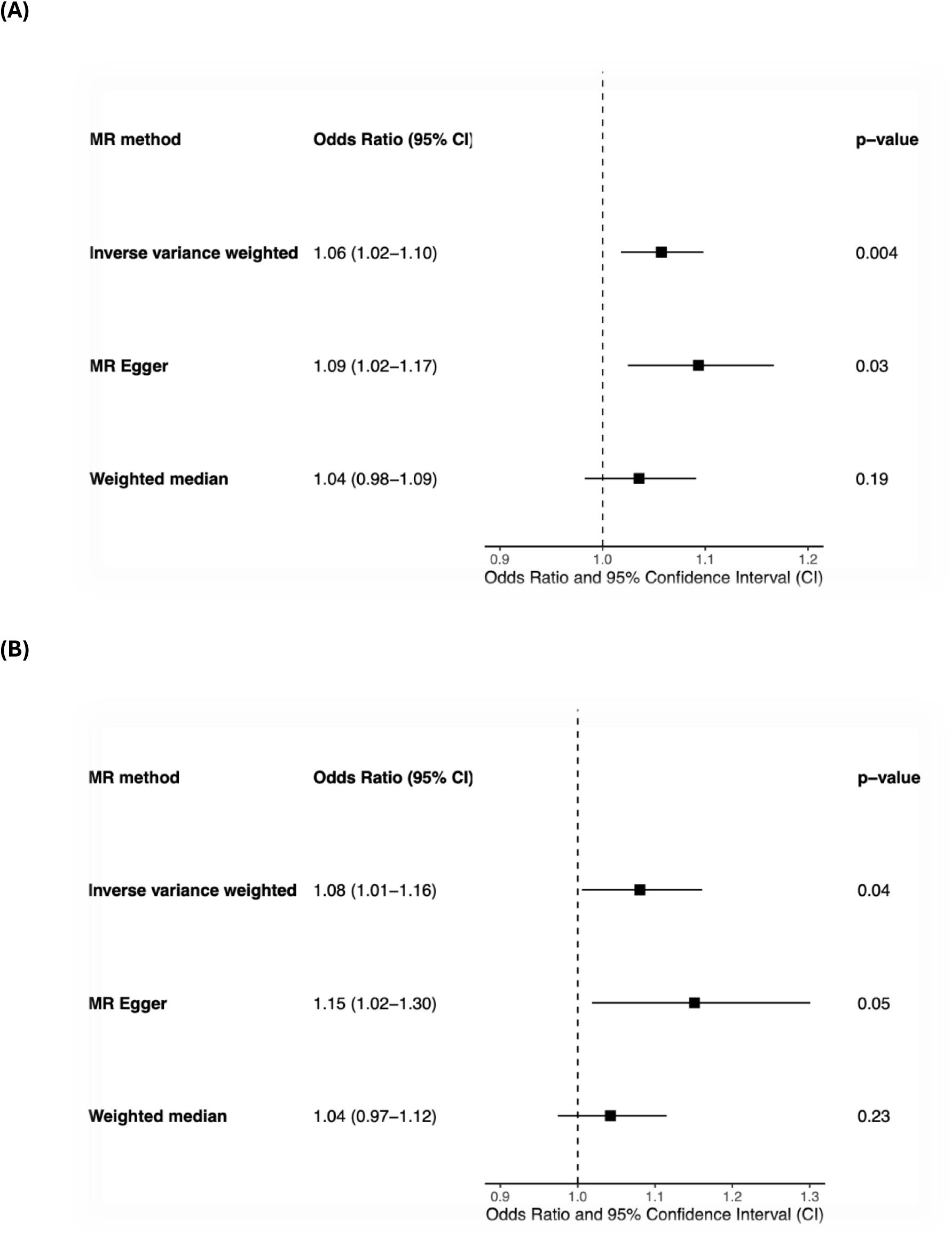
Mendelian randomisation (MR) sensitivity analysis results for the association between genetically predicted momp D antibody seropositivity level and (A) overall and (B) high-grade serous epithelial ovarian cancer risk. All odds ratios and confidence intervals are in units of per standard deviation increase in log-transformed antibody mean fluorescence intensity.

### Replication Mendelian randomisation and meta-analysis

We replicated our primary MR result for overall EOC risk using an independent GWAS data set that comprised the UK Biobank and FinnGen biobank combined, observing an association between genetically predicted elevated momp D antibody seropositivity level and increased EOC risk (OR 1.11 (95% CI 1.01—1.22; *p*=0.04; Figure 4), by the inverse variance weighted method. The point estimate of the effect size in the replication analysis was consistent between the inverse variance weighted method and the MR Egger and weighted median sensitivity analyses (Supplementary Tables 3 and 4). Fixed effect meta-analysis of the MR estimate from OCAC with that from the two biobanks yielded a combined OR of 1.06 (95% CI 1.03—1.10; *p*=0.0006; Figure 4).

**Figure 4:**
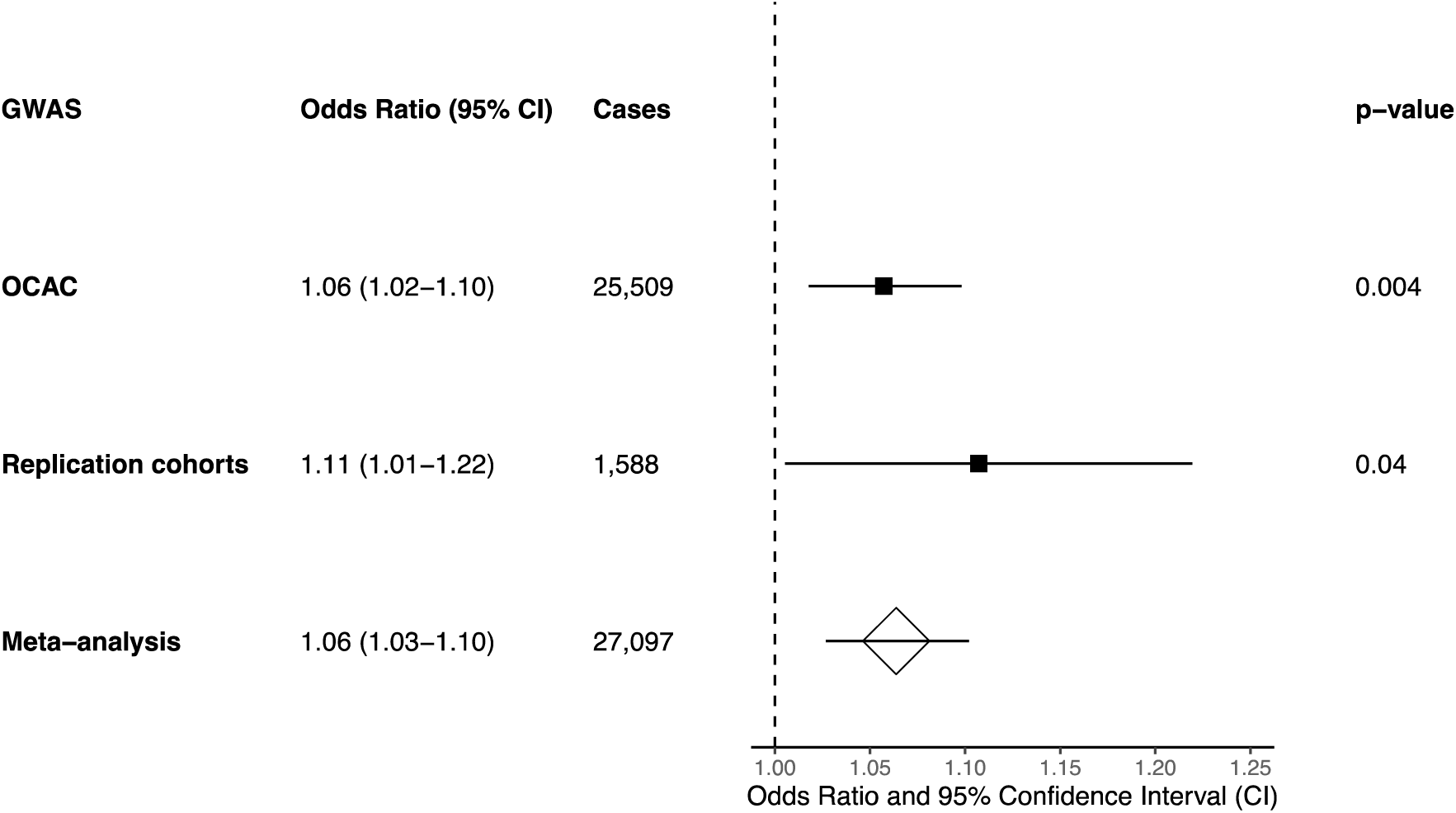
Replication Mendelian randomisation analysis (inverse variance weighted method) and meta-analysis results for the association between genetically predicted momp D antibody seropositivity level and ovarian cancer risk. The replication cohorts were UK Biobank and FinnGen combined. The meta-analysis combined the estimate from the replication cohorts with that from the Ovarian Cancer Association Consortium (OCAC) case-control studies. All odds ratios and confidence intervals are in units of per standard deviation increase in log-transformed antibody mean fluorescence intensity.

## Discussion

In this two-sample MR study, genetically predicted seropositivity to the *C. trachomatis* major outer membrane protein (momp) D antibody was causally associated with overall and high-grade serous EOC. This association with overall EOC remained when replicated in an independent population. There was no evidence of an association existing with other EOC histotypes.

This finding is partially consistent with a number of published observational studies investigating the relationship between exposure to STIs and EOC. A UK-based prospective analysis by Idahl *et al* using the European Prospective Investigation into Cancer and Nutrition (EPIC) cohort, including 791 EOC cases and 1669 controls, found *C. trachomatis* infection history (assessed by antibodies to chlamydia heat shock protein 60 (cHSP60-1)) was associated with higher risk of overall (1.36 [1.13-1.64]) and high-grade serous (1.44 [1.12-1.85]) EOC.^5^ Similarly, Fortner *et al* found *C. trachomatis* Pg3p antibody seropositivity was associated with higher risk of ovarian cancer overall (RR = 2.07 [1.25–3.43]) using 337 cases from the Nurses’ Health Studies. Their results were similar for invasive and low-malignant potential tumours, and no associations were observed for other STIs.^3^ Trabert *et al* conducted two case-control studies of 224 and 160 EOC cases in Poland and the USA respectively, finding *C. trachomatis* Pg3p antibody seropositivity to be associated with double the risk of EOC after adjustment. Again, no significant associations with other STIs were observed.^4^ However, a Finnish prospective study of 484 cases by Skarga *et al* found no association between *C. trachomatis* Pgp3 seropositivity and EOC, although did between *M. genitalium* and mucinous EOC (RR, 1.66 [95% CI, 1.09-2.54].^14^ Two further case-control studies from Italy and Canada found no significant association between C. trachomatis and EOC.^17,18^

Interestingly, findings from observational studies exploring the association between PID and EOC are also mixed. Jonsson *et al* conducted a case-control study of 15 072 Swedish EOC cases, reporting associations with PID (OR of 1.39; 95% CI 1.17—1.66).^13^ Meanwhile, a prospective study of 5356 women in Denmark found no associations with PID (hazard ratio (HR), 1.05; 95% CI, 0.92—1.20).^16^ A prospective study of 454 Australian women found high-grade serous EOC was associated with previous PID (HR 1.47; 95% CI 1.04-2.07), but not endometriosis or infertility.^15^ Conversely, a study of 600 African-American women found PID to not be associated with EOC, but endometriosis was.^29^ A further pooled analysis of 13 case-control studies concluded the relationship to be uncertain and liable to uncontrolled confounding.^30^

Some small-scale non-observational studies support the role of *C. trachomatis* in EOC carcinogenesis, whereby *C. trachomatis* DNA has been detected in 84% of high-grade serous and 17% of invasive EOC.^7,19^ Although limited by sample size, these studies may elucidate potential causative mechanisms underlying the observed association. Due to the high prevalence of asymptomatic *C. trachomatis* infections, exact incidence is unclear. It has been suggested that without treatment, approximately half of *C. trachomatis* infected women will develop chronic infection.^31^ Therefore, *C. trachomatis* infections often persist, and the link between chronic infection, inflammation and cancer has been extensively described.^32^ Additionally, *C. trachomatis* bacteria have the capability to enter a viable but non-replicative persistent state, effectively evading the immune response of the host cell.^33,34^ During this persistent state, the bacteria increase the production of cHSP60-1, an anti-apoptotic protein, while decreasing the synthesis of structural and membrane proteins.^34^ *C. trachomatis* can inhibit the release of mitochondrial cytochrome C and caspase 3, thus enabling the infected cell to evade intrinsic apoptosis.^35^ This ability to resist apoptosis prolongs the survival of the infected host cell and allows potentially DNA-damaged cells to persist, heightening the risk of STIC lesions and carcinogenesis.

This study has several strengths. First, we utilised MR to assess the causal association of genetic susceptibility to *C. trachomatis* with EOC, which can provide reliable evidence on the effect of modifiable risk factors for disease and overcomes some limitations of traditional observational epidemiology – such as study feasibility, ethical constraints, and confounding. Second, there were ten momp D seropositivity-associated lead SNPs, allowing us to conduct multiple sensitivity analyses to ensure robustness of our findings and determine the absence of horizonal pleiotropy. Third, our findings of an association have been further strengthened by positive associations holding when replicated using an independent Global biobank population. Finally, we used meta-analysis of our findings to give an overall effect estimate which may be useful to inform clinical or public health decision making.

Nonetheless, there are limitations. First, MR studies rely on a key assumption that the genetic instruments only act on the outcome via the exposure. Although this study yielded an insignificant MR-Egger intercept test, the possibility of horizontal pleiotropy cannot be completely ruled out. Third, the weighted median, simple mode, and weighted mode methods resulted in insignificant association in all the analyses, most likely attributable to the lower statistical power with reference to other MR methods.^36^ For the primary analysis using the OCAC GWAS, case numbers among the less common histotypes were limited, potentially underpowering results. For the replication analyses using the Global biobanks GWAS, outcome data combined all EOC histotypes together – both serous and non-serous – for which the pathology and epidemiology are known to differ.^7^ We used *C. trachomatis* momp D-associated lead SNPs as our genetically predicted exposure; Butler-Laporte et al found momp D to have a relatively high genomic inflation factor of 1.24.^21^ This questions whether all the SNPs identified show a true association with the phenotype (i.e. momp D seropositivity).^37^ However, the corrected inflation factor (λ_1000_) is 1.10 when the population size is considered, which is generally considered acceptable for GWAS.^38^ Furthermore, momp D is not the gold standard serological test for *C. trachomatis*. Although momp D antibody specificity is directly comparable to that of the Pgp3 antibody, Pgp3 is recognised as the gold standard as it is more sensitive and may be associated with longer persistence than momp D.^39^ However, Pgp3 was only associated with one SNP, unlike momp D which was associated with eleven, therefore it was not possible to use this as our study exposure given inability to perform sensitivity analyses. Both Pgp3 and momp D antibodies are general markers of *C. trachomatis* infection, and do not necessarily indicate persistent, untreated *C. trachomatis* infections which we postulate are causally associated with EOC. Many people infected with *C. trachomatis* in their lifetime do not develop chronic infection due to natural immunity or treatment, and it is unlikely these individuals will develop any long-term consequences – such as EOC. cHSP60-1, when produced by C. trachomatis bacteria, is associated with a state of persistent, untreated chlamydia infection and its sequelae – including evasion of immune defence, inhibition of cell apoptosis, and survival of a DNA-damaged cell. Therefore, it is proposed that cHSP60-1 production is a potential pathogenic mechanism by which *C. trachomatis* could cause EOC, thus positive serology for cHSP60-1 antibodies would be a more useful marker than momp D or Pgp3 to investigate whether an association with EOC exists.^5,7^ Unfortunately, no GWAS has yet included cHSP60-1 antibodies. It is also important to note that effect estimates, representing lifetime risk, may be somewhat underestimated due to prevalence of momp D seropositivity in individuals previously treated or naturally cleared of *C. trachomatis* infection.^40^ Finally, the GWAS meta-analysis from which the summary statistics were retrieved for this study only included Europeans and a small sample from Japan; therefore the generalisability to other populations are unknown.

This study, shedding light on the causative link between *C. trachomatis* and EOC, has several public health implications. Affecting approximately one in twenty women aged 15-24 years in the USA, *C. trachomatis* is highly prevalent and often silent.^41^ Our study provides compelling further reasons why accessible sexual health education and services are vital for short and long-term population health protection targets. The first phase one trial for a *C. trachomatis* vaccination was successfully completed in 2019, with others in pre-clinical development.^42,43^ Our study, highlighting this causative link, may further motivate stakeholders to continue supporting the development of *C. trachomatis* vaccines, with an aim to roll out in adolescence alongside HPV-vaccines.^43^ The causal role of *C. trachomatis* in the pathogenesis of other cancers, including anorectal and cervical, also requires further evaluation,^44–46^ as does EOC risk in the context of other STIs. *M. genitalium* is a small intracellular bacterium known to cause PID, and although less prevalent than C. trachomatis, it is also often asymptomatic, associated with PID, and frequently resistant to antibiotics. Observational studies have described associations with EOC, and with prevalence rising, it deserves attention.^3,5,14^ A study by Shanmughapriya *et al* suggested associations exist between intrauterine device (IUD) use and subsequent PID and EOC due to IUD colonisation by actinomycetes and disruption of the normal genitourinary flora.^47,48^ This is a further area requiring additional exploration, given the potential implications if found to be true. Finally, our study has highlighted STI exposure as an important aetiological aspect of EOC and therefore may have a role in cancer polygenic risk scores and stratified population screening strategies.^49^

In conclusion, this MR study confirms the causative link between *C. trachomatis* infection and overall and high-grade serous EOC. As a key modifiable risk factor for future serous EOC, primary prevention of *C. trachomatis* infection is a crucial public health target and may help reduce burden of EOC much like HPV-vaccination has for cervical cancer. Further studies into the causative role of other STIs and EOC, particularly *M. genitalium*, and *C. trachomatis* and other cancers, namely anorectal, may represent further opportunities to reduce cancer risk.

### Data availability

All data sets used in this study are publicly available at https://gwas.mrcieu.ac.uk/datasets/ with the following data set identifiers – OCAC ovarian cancer: ieu-a-1120 (overall EOC), ieu-a-1121 (high-grade serous EOC), ieu-a-1122 (low-grade serous EOC), ieu-a-1123 (invasive mucinous EOC), ieu-a-1124 (clear cell EOC), ieu-a-1125 (endometrioid EOC), ieu-a-1230 (serous low malignant potential EOC), ieu-a-1232 (mucinous low malignant potential EOC); UK Biobank + FinnGen ovarian cancer: ebi-a-GCST90018888; UK Biobank *Chlamydia trachomatis* momp D antibody seropositivity level: ebi-a-GCST90018888.

## Data Availability

All data sets used in this study are publicly available at https://gwas.mrcieu.ac.uk/datasets/

## Acknowledgements

We thank all participants in the OCAC studies and the UK Biobank and FinnGen cohorts.

## Author information

### Contributions

SPK conceived and designed the study. SLP analysed the data, prepared figures and tables, and wrote the initial draft of the manuscript. Both authors edited the initial draft of the mansucript and approved the final version.

## Ethics declarations

### Ethics approval and consent

This study is a secondary analysis of publicly available GWAS summary statistics data sets.

